# Inflammatory proteins as strong predictors of death in COVID-19 patients with coexisting neurological diseases

**DOI:** 10.1101/2024.02.27.24303438

**Authors:** Justyna Zielińska-Turek, Grzegorz Turek, Tomasz Lyson, Jan Gajewski, Mirosław Ząbek, Małgorzata Dorobek

**Author notes:** **Corresponding author:** Justyna Zielińska-Turek, MD, PhD; 137 Wołoska Str., 02-507 Warsaw, Poland, Wołoska 137, 02-507 Warsaw, Poland.

## Abstract

**Background:** Although many studies published so far on COVID-19 have examined its clinical prognosis, there is still no universal laboratory test that can assess the risk of a fatal outcome in patients with coexisting neurological diseases.

**Methods:** The plasma concentrations of C-reactive protein (CRP), procalcitonin (PCT), lactate dehydrogenase (LDH), ferritin, interleukin 6 (IL-6), D-dimers, and highly sensitive troponin I (hsTnI) were determined in a group of 400 consecutive in-patients with COVID-19 and concomitant neurological comorbidities.

**Results:** The median concentration levels of most of the inflammatory mediators/indicators, calculated for the whole group of patients, remained above the normal reference ranges, whereas the median concentrations of these substances were much higher in the sub-group of patients who died. Backward stepwise logistic regression confirmed the statistically significant predictors of death in a descending order of odds ratios as follows: LDH (3.8), ferritin (2.8), hsTnI (2.0), IL-6 (1.7), and age (1.01). A concentration of hsTnI > 64 ng/L appeared to constitute a strong predictor of an unfavorable prognosis. Patients who were treated with lopinavir and ritonavir, who required mechanical ventilation, and treatment with dexamethasone presented with significant increase in the concentrations of all the studied inflammatory proteins and increased odds ratio for death.

**Conclusion:** High plasma concentrations of pro-inflammatory proteins in patients suffering from COVID-19 and concomitant neurological diseases were associated with a more serious clinical course and an increased risk of death. The presence of these substances is worth monitoring as a valuable indicator of the current clinical condition of COVID-19 patients.

## Introduction

COVID-19 is a systemic disease that can develop as the result of an infection with the beta coronavirus SARS-CoV-2 [1,2,3]. The disease itself may be asymptomatic or may manifest as a severe condition with flashy inflammation and systemic coagulopathies, resulting in severe damage to the lungs, the cardiovascular system, and numerous other organs, including the brain. Respiratory symptoms predominate the clinical picture, ranging from a cough and dyspnea to interstitial pneumonia or acute respiratory distress syndrome [3,4,5,6].

The clinical course is often unpredictable and little is known about the factors that influence the acute deterioration of a patient’s clinical status. Many researchers have raised the issue of a “cytokine storm”—a mechanism responsible for the aggravation of symptoms and disease progression [7]. By activating the immune system, SARS-CoV-2 can stimulate the uncontrolled secretion of pro-inflammatory cytokines: mainly interleukin 6 (IL-6), but also other chemokines, TNF-α, and interferons [8,9]. Such an abnormal systemic inflammatory response can lead directly to ARDS and multiple organ failure, most often due to disseminated thrombosis [10-12].

IL-6 is a highly pleiotropic cytokine, and in excess, it can disrupt almost all body tissues. In particular, it causes the liver to produce acute-phase proteins and enzymes: C-reactive protein (CRP), procalcitonin (PCT), ferritin, and lactate dehydrogenase (LDH), among others. IL-6 is also known for its ability to signal in the central nervous system, eventually provoking a fever and malaise with an altered consciousness, nausea, arthralgia, and muscle pains.

COVID-19 can present with a broad array of clinical syndromes varying from a mild, flu-like form to a dramatic, severe illness with an unfavorable prognosis. A question then arises as to the factors that can predispose a patient to the serious form of the disease. This problem becomes even more serious when applied to a population of patients affected with a comorbidity due to pre- or/and coexisting neurological diseases. Assuming that the severe course is associated with excessive cytokine activity, one would need to look for a set of biochemical parameters that can reliably herald the presence and mirror the intensity of a cytokine storm with all its undesirable effects. In addition to CRP and ferritin, increases in D-dimers, procalcitonin, and highly sensitive troponin I (hsTnI) have been cited as participating in this dramatic result.

Currently, there is no universal set of laboratory tests that can determine the prognosis in a patient with the coronavirus disease. The aim of this study was to determine the relationship between concentrations of inflammatory proteins (determined upon the patient’s admission), the changes in radiographs of the lungs, and the risk of death in patients with COVID-19 who also suffer from pre- or/and coexisting neurological diseases.

## Materials and Methods

This prospective study was conducted by the biggest Polish neurological department dealing with COVID-19 during the world pandemic—the Central Clinical Hospital of the Ministry of Internal Affairs and Administration in Warsaw. The entire study was carried out in accordance with the tenets of the Declaration of Helsinki (as revised in 2013) and was approved by the local ethics committee of the Central Clinical Hospital in Warsaw (85/2020). All the involved patients gave their informed consent.

Data were collected from March 1 2020 to March 31 2021. The study included 400 adult in-patients (age of ≥18 years; 207 women, mean age of 69.1±17.6; and 193 men, mean age of 65.2±15.5) hospitalized due to a variety of neurological conditions and concomitant COVID-19 (Tables S1 and S2). Among them were 28 patients affected by more than one neurological disease. A SARS-CoV-2 infection was confirmed at the time of admission using real-time PCR (RT-PCR) on a swab from the nasopharynx. The PCR test was carried out by a specially trained nurse, and a neurological diagnosis was made by a specialist in neurology, supported with MR, CT, and Doppler US imaging of the head and—when necessary—the chest. Upon admission, the patients were also tested for the presence of influenza viruses A and B as well as for the respiratory syncytial virus. Individuals with positive findings were excluded from the study. All patients with procalcitonin > 0.5 ng/mL, with typical pattern of bacterial lung infection in chest computer tomography were suspected for potential concomitant bacterial infection and excluded from database. Patients with potential lowered immunity (e.g. during long term antibiotic or immunosuppressive treatment) were suspected for potential fungal infection and also excluded from database.

The general condition of the patients—in particular, their respiratory efficiency and consciousness—was assessed using the modified early warning score (MEWS) and the Glasgow coma scale (GCS). The NIHSS (National Institute of Health stroke scale) was used for scaling the neurological deficits. The patients’ cognitive functions were assessed with the mini-mental state examination (MMSE).

A set of laboratory investigations was performed no later than 6 hours after admission. In addition to a complete blood count, it included an assessment of the plasma concentrations of C-reactive protein (CRP), procalcitonin (PCT), lactate dehydrogenase (LDH), ferritin, IL-6, D-dimers, and highly sensitive troponin I (hsTnI). The concentration of plasma D-dimers (normal range: up to 500 µg/L) was determined using the VIDAS D-dimer test (bioMérieux). The troponin I concentration levels were assessed with the hsTnI assay (Abbott Diagnostics). Median values and ranges of the concentration levels were calculated for the sub-group of survivors and for the patients who finally died (Table S1, Figure S1), as well as separately for each specific neurological disorder (Table S2).

### Statistical Analysis

The distributions of the studied variables were estimated with the Kolmogorov–Smirnov test and appeared to be significantly right-skewed. For this reason, the median as well as the lower/upper quartile values were used to represent the distributions and non-parametric tests were used for testing the differences between the groups: the Mann–Whitney test with a continuity correction and a chi-squared test with an estimation of the effect size using Cramer’s φ. *P*-values were assessed with the Bonferroni correction. The Glass rank-biserial correlation (R_G_) was used to assess the effect sizes. Backward stepwise logistic regression was used to determine the probability of death depending on the set of individual laboratory parameters, the neurological diagnosis, and the treatment modality (Table S3). The effect of concomitant neurological diseases on the risk of death was tested with a log-linear analysis.

## Results

### Demographic and Baseline Characteristics

The neurological assessment was performed at the time of admission and—for the survivors—upon discharge. The mean scores for the entire group in subsequent clinical scales upon admission and discharge were as follows: MEWS, 3.2 ± 2.5 and 1.5±1.1; GCS, 11.8±7.6 and 14.1±5.3; NIHSS, 14.2±5.2 and 10.8±11.1; and MMSE, 17.3±5.4 and 24.1±7.8. Out of 400 patients, the outcomes in 95 individuals were fatal.

### Clinical Characteristics of the Study Group

The most common neurological condition was ischemic stroke, which occurred in 201 patients (50.2%)—106 men (mean age of 68.6 years ±13.7) and 95 women (mean age of 73.2 years ±14.8). Out of them, 18 underwent thrombolysis and 15 were provided with a mechanical thrombectomy. The other conditions included dementia (7.25%), epilepsy (7.25%), CNS tumors (5%), hemorrhagic stroke (4.5%), diseases of the neuromuscular junction (2%), demyelinating diseases (3%), and other conditions (20.8%) (Table S2). More than one neurological condition (overlapping diseases) was diagnosed in 28 patients (Table S4).

The most common coexisting non-neurological disorder was arterial hypertension (205 patients), followed by diabetes (97 patients), atrial fibrillation (70 patients), and a neoplastic tumor (31 patients). Renal failure (29 patients), a nicotine addiction (21 patients), significant obesity (18 patients), asthma (13 patients), and an alcohol addiction (13 patients) were noted less frequently.

Thrombosis/an embolism was found to be a main factor in 41 out of the 201 patients with ischemic stroke, diagnosed with angio-CT (computer tomography) imaging. A cardiogenic mechanism of stroke was suspected in 61 out of the 201 patients (those presenting with arrhythmias or atrial fibrillation). Forty-nine patients presented with coexisting hypertension, dyslipidemia, type 2 diabetes, or an addiction to nicotine. The remaining patients with ischemic stroke presented with end-stage renal failure and/or disseminated neoplastic disease. Dementia was the second most frequent condition in the whole study group, and the patients with dementia were older in relation to the general study population (mean of 82 ± 7.7 years and range of 67-99 years vs. mean of 67±16.7 years and range of 19-99 years, respectively; *p*<0.05), and all of them had a number of coexisting comorbidities that may have negatively influenced the prognosis.

The most common symptom present in our patients was a fever (25%), followed by dyspnea (23%), a cough (12%), myalgia (10%), dizziness (7%), a headache (9%), vomiting (7%), changes in taste or smell (3%), diarrhea (4%), and constipation (2%). Chest computer tomography (CT) revealed interstitial densities, a crazy paving pattern, ground-glass opacities, and exudate—abnormalities typically found in patients with advanced forms of COVID-19. Nevertheless, CT pictures of the chest with no discernible pathology were also relatively common.

### Treatment and Outcomes

The treatment of patients with advanced COVID-19 focused on mechanical support of the respiratory function. Out of 400 patients, as many as 70 received high-flow oxygen therapy and twenty-one required mechanical ventilation. Unfortunately, the mortality amounted to 90% for the ventilated patients, in spite of their treatment in the intensive care unit. Altogether, 95 patients out of the studied 400 died. The mortality in the group of 28 patients with more than one concomitant neurological condition did not differ from the mortality in the general group, as tested with a log-linear analysis. In addition, the concentration levels of virtually all the studied inflammatory proteins did not differ to any statistically significant degree from those detected in the general group (Mann–Whitney U test).

In the initial period of the pandemic, chloroquine was used in more than half of the patients (57 persons). Azithromycin (35%) and/or ceftriaxone (41%) were used as an adjuvant therapy in patients with interstitial pneumonia. Nine patients who were resistant to treatment with chloroquine received antiviral therapy with lopinavir and ritonavir. Unfortunately, seven out of the nine patients died, and additionally, three of them demonstrated laboratory signs of pancreatitis. With the emerging reports on the effectiveness of remdesivir, our protocol changed to include treatment with this drug, either alone or in combination with convalescent plasma. Both media were applied during the first 7 days since the onset of symptoms, and were then replaced with tocilizumab for the next week.

Among the group of 40 patients treated with remdesivir, only four deaths occurred, and—of note—the hospitalization time of the survivors was shorter by 3.3 days, on average, than that of the survivors who were not treated with this antiviral medication. The results of the treatment with convalescent plasma did not differ from the results obtained with remdesivir. There were four deaths among 20 patients and the hospitalization time of those who survived was, on average, 4.1 days shorter than that of the survivors who were not treated under this regime in the initial phase of the pandemic. Altogether, 40 patients were treated with remdesivir, 20 were treated with convalescent plasma, and only 2 patients received tocilizumab. Additionally, 36% of the patients were supplemented with dexamethasone during their oxygen therapy.

Logistic regression of the variables relating to the diagnosis and treatment modalities demonstrated increased odds ratios of death for the patients who were treated with lopinavir and ritonavir (OR: 14.8), those who required mechanical ventilation (OR: 10.2), and those treated with dexamethasone (OR: 2.1, Table S3).

### Concentration of Inflammatory Mediators

The median concentration levels of CRP, ferritin, D-dimers, and lactate dehydrogenase (LDH), calculated for the whole group of patients, remained above the normal reference levels. The overall median concentrations of IL-6, lymphocytes, procalcitonin, and hsTnI remained within the normal reference ranges (Table S1).

In the three most frequent neurological conditions concomitant with COVID-19, the concentrations of inflammatory mediators were as follows. The concentrations of D-dimers in patients with ischemic stroke were more than three times higher in comparison to the normal range (1730 mg/L vs. <500 mg/L; Table S2). As for the remaining markers in this sub-group of patients, the procalcitonin concentration remained within the normal reference level, whereas the concentrations of C-reactive protein and ferritin increased 3-fold (Table S2). In patients with dementia, the procalcitonin level remained within the normal reference range, whereas the C-reactive protein concentration increased 6-fold and the concentration of ferritin increased 4-fold (Table S2). In patients with epilepsy, the C-reactive protein level increased 4-fold, but IL-6 and hsTnI were also significantly increased. The concentrations of inflammatory mediators in our clinical sub-groups of patients are shown in Table S2.

The concentration levels of all the tested inflammatory proteins turned out to be significantly higher in patients who required mechanical ventilation (p<0.05), and in those who were treated with dexamethasone (p<0.001) or lopinavir and ritonavir (p<0.05), compared to those who did not require such vigorous therapy (Mann–Whitney test).

#### Patients Who Eventually Died vs. Survivors

The largest differences between the fatality group and the convalescent group concerned the concentrations of ferritin, interleukin 6, LDH, and troponin I (large-effect eta2> 0.14, Table S1, Figure S1). The plasma concentrations of all the above substances were significantly lower in the convalescent patients than in the deceased patients. The median concentration of hsTnI (an agent often associated with heart biology) was significantly higher in the deceased patients in comparison to the survivors. At the same time, the concentration of this substance significantly increased with an advancing age of our patients. An initial hsTnI concentration > 64 ng/L proved to be the strongest predictor of death in a model computed with backward stepwise logistic regression. This model was able to predict death with a sensitivity of 62% and a specificity of 95%. The best model, which included age and the logarithms of inflammatory parameter concentrations, is shown in Table S3.

Compared to the convalescent group, the median age of the patients who died was significantly higher (64.2 ± 16.9 years vs. 76.8 ± 11.4 years, Z=6.7, *p*<0.0001, R_G_=0.45). There was no effect of gender on the concentration of all the analyzed inflammatory proteins in either outcome group (all effects were considered trivial; R_G_<0.1). The convalescents comprised 79% women and 73% men, and the frequency difference was not significant: χ^2^=1.48, *p*=0.22, and φ=0.061.

In the sub-group of survivors, all the concentration levels were lower compared to the levels of the entire study group, whereas in the group of deceased patients, they were significantly higher than in the general group. In the group of convalescents, only the median values of procalcitonin and hsTnI met the upper range of normal values for the healthy population (0.08 ng/mL and 7.7 pg/mL, respectively), whereas the median values of the other parameters were slightly above the normal reference range (Table S1).

## Discussion

To date, there are no universal laboratory tests that are useful for predicting the prognosis of patients with the coronavirus disease. It is, however, well known that a severe SARS-CoV-2 infection affects the immune system, releasing pro-inflammatory cytokines in an uncontrolled manner (a phenomenon known as a “cytokine storm”). Based on the above premise, we attempted to determine the relationship between the concentrations of inflammatory proteins and the risk of death in patients with COVID-19 and coexisting neurological diseases.

The set of investigated cytokines included IL-6, which is over-released to recruit immune cells against the virus [8]. The abnormal recruitment of leukocytes affects numerous organs, especially the lungs, leading to acute respiratory distress syndrome (ARDS) [13,15,16]. Zhou et al. documented that, besides IL-6, D-dimers, ferritin, and lactate dehydrogenase may also participate in the cytokine storm, thus increasing the risk of death [17]. Zeng et al. confirmed that increased levels of IL-6 are correlated with the rate of mortality in COVID-19 patients [18]. Excessive IL-6 secretion may also contribute to a reduction in the number of lymphocytes. Another adverse effect of IL-6 is the abnormal induction of the coagulation cascade, which may also contribute to the sudden deterioration of a patient’s clinical condition [19-28].

Taking into account the important role of excess IL-6 in the development of dangerous biological reactions, the determination of this cytokine could be used for monitoring COVID-19 patients in order to predict the deterioration of their clinical status. Several IL-6 concentration levels have been established and proposed as diagnostic thresholds for identifying an increased risk of clinical deterioration or death. Elshazli et al. and Guirao et al. proposed the thresholds of 22.9 pg/ml and 34.9 pg/ml, respectively, and both were used to predict the need to transfer a patient to the intensive care unit (ICU) [29,30]. Riveiro-Barciela et al. stated that an IL-6 concentration amounting to 64 pg/ml may predict the need for high-flow oxygen therapy (hazard ratio of 18) [31]. Mandel et al. established that an IL-6 level of 163.4 pg/ml was a strong predictor of death, with a sensitivity of 91.7% and a specificity of 57.6% [32]. In our study, Il-6 also appeared to be a strong (but not the strongest, among other pro-inflammatory proteins) predictor of death. The odds ratio for this dramatic effect, calculated using backward stepwise logistic regression, amounted to 1.67. The difference between the concentration of IL-6 in the convalescent group and in the deceased group was statistically highly significant (10.1 pg/mL vs. 37.65 pg/mL).

The troponin (hsTnI) levels were assessed in every patient included in this study group, regardless of whether signs of acute coronary syndrome were present, because chest pain (which usually accompanies COVID-19) may mimic heart failure. An increased concentration of hsTnI is a reliable secondary marker of cellular damage and, thus, may also be present in viral and bacterial infections, sepsis, vascular diseases of the central nervous system, acute respiratory distress syndrome (ARDS), and renal insufficiency [17]. Zhou et al. showed that the concentration of hsTnI was elevated in patients with COVID-19 in an already-advanced or progressing disease, especially in those who eventually died [17]. Our results are consistent with this and support hsTnI as a strong predictor of death. An initial concentration of hsTnI above 64 ng/L was a strong predictor of death in our study group, with the odds ratio amounting to 2.02. Notably, the hsTnI concentration was higher not only in those patients who eventually died, but also in individuals of an advanced age.

Lippi et al. showed that assessments of the procalcitonin concentration may be useful in determining the risks of COVID-19 complications, clinical deterioration, and death [15]. In our study, the procalcitonin concentrations in the convalescents met the upper limit of the normal reference range for the population (0.08 ng/mL), whereas in the group of patients who finally died, it was two times higher. However, contrary to the results of Lippi et al., our results suggest that procalcitonin cannot serve as a reliable predictor of death (*p*>0.05 in backward stepwise logistic regression). Interesting results were published by Rodelo et al., who performed a retrospective analysis on patients with a suspected infection/sepsis and found that the median levels of D-dimers in non-survivors increased to 2489 ng/mL. The odds ratio of death for this level of D-dimers amounted to 3.03, with a 95% confidence interval between 1.38 and 6.62 [33]. In turn, regarding D-dimers, the concentration of this substance was found to be increased in the patients who finally died, but no statistically significant relationship between higher levels of D-dimers and the risk of death was demonstrated using a backward stepwise logistic regression analysis (*p*>0.05).

Lactate dehydrogenase (LDH) is an enzyme involved in energy conversion, and is, therefore, present in almost all the cells in the body. Tests for determining the concentration of LDH in the blood are commonly used to monitor tissue damage in a wide range of disorders, including liver disease, interstitial lung disease, and the illness induced by SARS-CoV-2. LDH has also been identified as an important biomarker of the activity and severity of idiopathic pulmonary fibrosis. It can be concluded that the dramatic increase in the LDH concentration that we observed in those patients who finally died can be regarded as an important prognostic marker, mirroring lung injury. Such an opinion is also supported by the data provided by Bartziokas et al., who found that the concentration of LDH increased in critically ill COVID-19 patients with the advancement of the disease and with the extent of lung injury [34]. These authors also demonstrated that an increased plasma LDH concentration is closely related to COVID-19 mortality. In addition, in our group of patients, a high level of LDH was the strongest predictor of death, with an odds ratio of 3.792.

Another protein known to influence the severity of COVID-19 is ferritin. Zhou et al. showed that individuals with advanced COVID-19 also had an elevated level of serum ferritin, and additionally, the serum ferritin was significantly higher in the “very severe” form of COVID-19 in comparison to merely “severe” cases of COVID-19: 1006.2 ng/ml (IQR: 408.3-1988.3) vs. 291.1 ng/ml (IQR: 102.10-648.42), respectively [35]. It was revealed by Chen et al. that, in patients who finally died of COVID-19, the ferritin levels were high upon hospital admission and throughout their hospital stay. The median values of the serum ferritin concentration of these patients after day 16 of hospitalization exceeded the upper limit of detection in these patients, suggesting that the ferritin levels increased non-stop [36]. Tao et al. analyzed the clinical characteristics of 99 patients, and among them, 63 had serum ferritin levels well above the normal range. An analysis of the peripheral blood of 69 patients with severe COVID-19 revealed elevated levels of ferritin compared to patients with non-severe disease. The authors concluded that serum ferritin levels are closely related to the severity of COVID-19 [37]. Our study supports this conclusion, as the serum levels of ferritin were significantly higher in those patients who subsequently died than in the survivors. An abnormally high serum concentration of this protein appeared to be a strong predictor of death, as demonstrated with backward stepwise logistic regression (odds ratio of 2.81).

An advanced age is also a known predictor of COVID-19 severity. Nevertheless, our findings demonstrated only a weak impact of age on the risk of death, with the odds ratio only slightly exceeding 1. This somewhat-unexpected result can most likely be explained by the fact that our study population had a significant burden of pre-existing and coexisting conditions, including diabetes, cardiovascular disease, chronic kidney disease, hypertension, obesity, lifestyle issues, etc., which may have increased the mortality independently of age [38-41]. In summary, a COVID-19 infection in patients with pre-existing or coexisting comorbidities carries a high risk of mortality, multi-organ dysfunction with respiratory failure, encephalopathy, acute heart damage, renal failure, and damage to other organs [35]. Guo et al. constructed a score named MuLBSTA (multilobular infiltrates, lymphocytes ≤0.8 × 109 / L, bacterial infection, smoking, hypertension, and age ≥60 years) that may help predict the outcomes in patients with COVID-19 [39].

Interestingly, the risk of death appeared to differ between the sub-groups of our patients, i.e., those affected with ischemic stroke, epilepsy, dementia, or other conditions. For each of the mentioned conditions, COVID-19 may change the risk of death in a different manner. The highest risk of death—approximately 52%—was noted among the patients with dementia. However, the median age in this group was the highest (amounting to 82 years), and the patients presented with an abundance of coexisting diseases. On the other hand, the group of patients with epilepsy, which was featured with a relatively low risk of death, were, on average, significantly younger (median age of only 62.9 years) than those with dementia. Few deaths occurred in this group of patients, mainly due to an uncontrollable epileptic state.

The treatment methods for our 400 patients were, of course, varied depending on their individual clinical condition. As much as 70 patients received high-flow oxygen therapy and 21 required mechanical ventilation. The mortality in the latter group amounted to 90%, and multivariable logistic regression only confirmed that the need for mechanical ventilation significantly increased the risk of death (odds ratio of 10.2). Notably, in that group of patients, all the tested inflammatory proteins were significantly higher (*p*<0.05) when compared to those who did not require mechanical ventilation. This result indicates that the need for artificial ventilation and a high concentration of cytokines upon admission show an inter-relationship, supporting our thesis on the prognostic importance of cytokine determination in neurological patients infected with COVID-19.

On the other hand, pro-inflammatory cytokine determination may also play an important role in predicting the need for mechanical ventilation during hospitalization. In a study on 1042 patients hospitalized due to a COVID-19 infection, Nicholson et al. showed that the mortality rate of mechanically ventilated patients increased with age: in the youngest (<35 years), it was close to 0%, whereas in the oldest (≥75 years), it amounted to 75% [42]. Interestingly, we did not find any significant relationship between age and the risk of death of mechanically ventilated patients in our study.

At the beginning of the pandemic, chloroquine was used frequently by our institution, in more than in half of the patients treated during that period. However, after publications questioned its role in the treatment of COVID-19, chloroquine was withdrawn in favor of other therapeutic agents [43]. In light of the above, it is somewhat surprising that no influence of chloroquine therapy on the risk of death was found in our group of patients according to multivariable logistic regression. In addition, the therapeutic use of azithromycin and/or ceftriaxone (which were used as an adjuvant therapy in patients with interstitial pneumonia) did not appear to interfere with the rate of death. Nevertheless, treating COVID-19 with lopinavir and ritonavir did increase the odds ratio of death up to 14.2, unlike the drugs mentioned above. These two drugs were used in patients who were resistant to treatment with chloroquine, and interestingly, that sub-group had significantly higher concentrations of inflammatory cytokines at the time of admission than the other patients from our clinical series. Nevertheless, the importance of that finding may be limited due to the small number of patients treated with lopinavir and ritonavir.

Meini et al. addressed the use of lopinavir and ritonavir in COVID-19 therapy in their review published in the *Journal of Clinical Medicine*; they stated that the available evidence on this subject is scarce and of low quality. The recommendations issued for COVID-19 vary, from positions clearly against the use of lopinavir and ritonavir to indications of other drugs that are more favorable. Nevertheless, the authors express their opinion that lopinavir and ritonavir should not be abandoned in the treatment of COVID-19, in spite of the sometimes-controversial results of clinical trials [44]. Along with further reports on the effectiveness of remdesivir, our protocol changed in favor of treatment with this drug, alone or in combination with convalescent plasma, followed by tocilizumab. The results of the present study demonstrated that the risk of death did not increase, as assessed with multivariable logistic regression. In turn, Beigel et al. proved that remdesivir was superior to a placebo in shortening the time to recovery in adult patients who were hospitalized due to COVID-19, with evidence of a reduced respiratory tract infection [45].

In 36% of our patients, oxygen therapy was supplemented with dexamethasone, and the odds ratio of death in this group increased to 2.1 in relation to the patients who did not require this kind of therapy. Similarly to the situation with the ritonavir-treated group, the concentrations of inflammatory mediators seen upon admission also appeared to be significantly higher in the patients who needed therapy with dexamethasone compared to those who were effectively treated without the need for dexamethasone. In line with our results, Ahmed et al. stated that the liberal use of corticosteroids is not advocated, as high doses of these drugs can cause more harm than good [46]. On the other hand, in a thorough study presented by the RECOVERY Collaborative Group (*The New England Journal of Medicine 2020*), it was shown that the 28-day mortality of 2104 in-patients with COVID-19 was improved if invasive mechanical ventilation or oxygen therapy was supported with dexamethasone [47]. This discrepancy in the results indicates that the exact role of corticosteroids in COVID-19 therapy requires further clarification.

### Limitations of the Study

Although our study group was rather large, the number of patients with overlapping neurological diseases (i.e., more than one) was relatively narrow, including only 28 subjects. Therefore, our conclusion regarding a lack of specificity for these patients in relation to the whole group (especially regarding morbidity) may, in the future, need upgrading after obtaining a larger representation of COVID-19 patients with multiple neurological comorbidities.

Another limitation relates to the high proportion of patients affected by other, non-neurological diseases in the study group. Conditions such as atrial fibrillation, renal failure, neoplasms located outside the CNS, etc., may have influenced the expression of the studied inflammatory proteins. On the other hand, such a situation is rather naturally expected, and any future study on a hypothetical population of patients with COVID-19 that are free of concomitant diseases would hardly be possible.

Within the framework of this study, a set of inflammatory proteins was assessed only upon admission to serve as a potential prognostic factor to predict the outcome of COVID-19 infections. For this reason, it is difficult to answer the question of whether the influence of their increased concentration on higher odds ratios of death was direct or indirect (by requiring specific treatment modalities, such as artificial ventilation, antiviral therapy, and/or corticosteroids).

## Conclusions

This study demonstrated a positive correlation between the concentrations of inflammatory proteins and the severity of the course of the COVID-19 disease, including a direct relationship between significantly increased concentrations of these proteins and the risk of death.

Patients who were treated with lopinavir and ritonavir, who required mechanical ventilation, or who required additional treatment with dexamethasone presented with both a statistically significant increase in the concentrations of all the studied inflammatory proteins and an increased odds ratio for death.

The group of concentrations of IL-6, LDH, ferritin, and hsTnI is proposed as a tool for monitoring COVID-19 patients with pre- or/and coexisting neurological diseases. An initial hsTnI concentration > 64 ng/L was a strong predictor of an unfavorable outcome.

## Data Availability

The data underlying the results presented in the study are available from corresponding author of the manuscript

## Acknowledgments

N/A

## Institutional Review Board Statement

The research involving human subjects complied with all the relevant national regulations and institutional policies; was conducted in accordance with the tenets of the Declaration of Helsinki (as revised in 2013); and was approved by the local ethics committee of the Central Clinical Hospital in Warsaw (85/2020).

## Informed Consent Statement

Informed consent was obtained from all the individuals included in this study.

## Data Availability Statement

The data presented in this study are available from the corresponding author upon request due to restrictions, e.g., privacy or ethical reasons.

---

**Figure S1.**
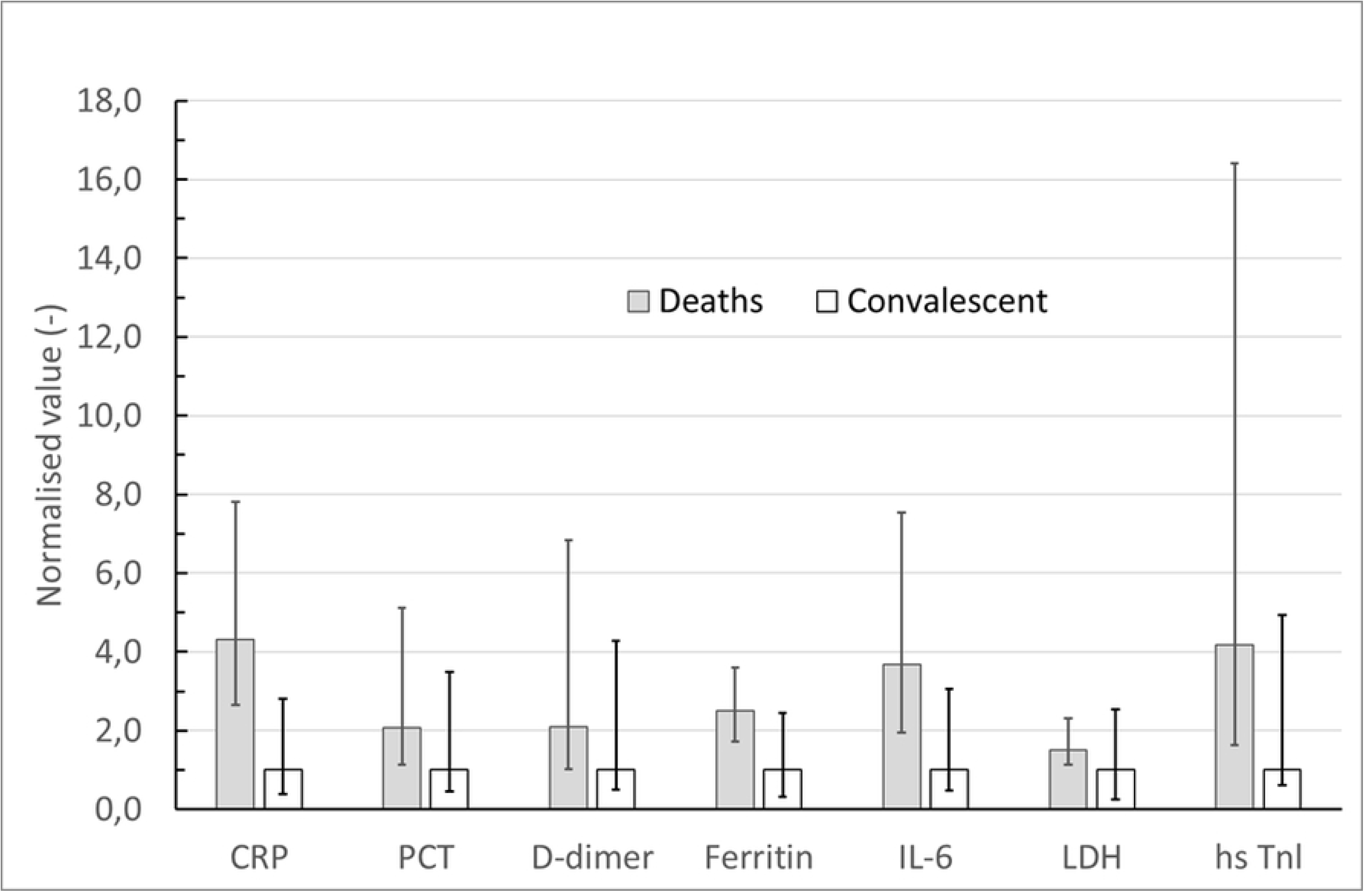
Concentration levels of inflammatory proteins in 400 COVID-19 patients in two groups: deaths and convalescents.

**Table S1.** Outcome and concentration levels of inflammatory proteins in 400 COVID-19 patients with coexisting neurological diseases.

|  | Total | Convalescents | Deaths | Mann–Whitney U test |  |  |  |
| --- | --- | --- | --- | --- | --- | --- | --- |
|  | Median | Median | Median | U | Z | *p*-value | R<sub>G</sub> |
|  | (lower Q–upper Q) | (lower Q–upper Q) | (lower Q–upper Q) |  |  |  |  |
| No. | 400 | 305 | 95 |  |  |  |  |
| Age (years) | 67.2 (19-99) | 64.2 (19-94) | 76.79 years (39-99) |  | 6.73 | 0.0000* | 0.457 |
| C-reactive protein (normal range: < 10 mg/L) | 26.55 (4.6-73.8) | 15 (2.7-56.7) | 64.7 (39.8-117.2) | 7031 | -7.52 | 0.0000* | 0.5115 |
| Procalcitonin (normal range: < 0.1 ng/mL) | 0.09 (0.05-0.19) | 0.08 (0.04-0.15) | 0.165 (0.09-0.41) | 7578 | -6.86 | 0.0000* | 0.4679 |
| D-dimers (normal range: < 500 ug/L) | 1230 (633-2476) | 1070 (526-2183) | 2234 (1095-7316) | 7980 | -6.06 | 0.0000* | 0.4193 |
| Ferritin (normal range: 24-336 ug/L) | 588 (297-1160) | 441 (268-870) | 1160 (774-1637) | 5540 | -7.94 | 0.0000* | 0.5640 |
| IL-6 (normal range: < 18ng/L) | 16.1 (5.02-39.1) | 10.1 (3.85-29.6) | 37.65 (20.55-76.15) | 5381 | -7.94 | 0.0000* | 0.5672 |
| LDH (normal range: 140-280 U/L) | 291 (205-390) | 262 (197-340.5) | 396 (297-608) | 6108 | -7.56 | 0.0000* | 0.5320 |
| Highly sensitive troponin I (normal range: < 15.6 pg/mL)<br>Female: < 15 pg/mL<br>Male: < 20 pg/mL | 12.3 (4.0-29.5) | 7.7 (3.1-20.4) | 43.8 (15.6-187.9) | 5503 | -8.99 | 0.0000* | 0.6136 |
| White blood cells (<10 tys./mL) | 7.41 (2.45-119.19) | 7.18 (2.45-119.19) | 9.44 (4.07-28.06) | 4965 | - | 0.0000* | - |

\* $p < 0.001$ shows statistical significance.

**Table S2.** Concentration levels of inflammatory proteins and number of deaths according to different diagnoses.

|  | Median (range) |  |  |  |  |  |
| --- | --- | --- | --- | --- | --- | --- |
|  | Ischemic stroke | Epilepsy | Dementia | CNS tumors | Hemorrhagic stroke | Others |
| No. | 201 | 29 | 29 | 20 | 18 | 103 |
| Age | 73 years<br>(38-98) | 62.9 years<br>(38-92) | 82.1 years<br>(67-99) | 71 years<br>(44-84) | 65 years<br>(23-84) | 63 years<br>(19-90) |
| Death (n) | 32 | 6 | 15 | 4 | 10 | 29 |
| C-reactive protein<br>(normal range: < 10 mg/L) | 28.5<br>(0.3-347.7) | 44.3<br>(1.6-196.1) | 44.2<br>(2.1-184.4) | 2<br>(0.2-177.2) | 17<br>(0.8-191.4) | 24.2<br>(0.2-404.6) |
| D-dimers (normal range: < 500 ng/mL) | 1730<br>(215-94437) | 1336<br>(221-23171) | 1129.5 (674-22580) | 1497.5<br>(149-8117) | 1626<br>(315-26426) | 1080<br>(181-28384) |
| IL-6 (normal range: < 18ng/mL) | 26.7<br>(2.03-3144) | 44.8<br>(1.5-3430) | 42.6<br>(2.49-243) | 4.94<br>(1.5-144) | 7.25<br>(0.9-81.3) | 10.1<br>(0.5-6493) |
| Procalcitonin (normal range: < 0.1 ng/mL) | 0.08<br>(0.008-226) | 0.15<br>(0.02-39.7) | 0.15<br>(0.07-2.85) | 0.07<br>(0.02-5.09) | 0.08<br>(0.04-0.37) | 0.1<br>(0.02-219) |
| Ferritin (normal range: 24-336 ug/L) | 503<br>(43-3721) | 951<br>(39-1403) | 795.5<br>(137-2473) | 560<br>(180-1650) | 390.5<br>(179-2159) | 562<br>(71-5515) |
| LDH (normal range: 140-280 U/L) | 303<br>(71-1024) | 345<br>(139-1180) | 285.5<br>(180-728) | 234<br>(187-338) | 257<br>(120-708) | 286<br>(117-1191) |
| Highly sensitive troponin I (normal range: < 15.6 pg/mL) | 15.2<br>(0.3-28287) | 22.65<br>(1.1-16148) | 19.1<br>(4.5-380.5) | 5.9<br>(1.1-45.1) | 11.15<br>(2.2-30.9) | 6.1<br>(0-275.5) |
| White blood cells<br>(normal range: < 10 tys/mL) | 7.63<br>(2.88-119.19) | 8.34<br>(3.5-14.68) | 7.53<br>(2.93-28.06) | 10.33<br>(4.7-19.85) | 7.58<br>(4.08-18.1) | 6.63<br>(2.87-26.34) |

**Table S3.** Statistically significant predictors of death in the group of 400 patients with COVID-19 and neurological comorbidities.

|  | Odds ratio | Confidence interval (CI) | *P* |
| --- | --- | --- | --- |
| Age | 1.066 | 1.03-1.10 | 0.0003* |
| Lopinavir and ritonavir | 14.8 | 2.90-75.3 | 0.0012* |
| Mechanical ventilation | 10.2 | 3.40-30.8 | 0.0000* |
| Dexamethasone | 2.1 | 1.29-3.52 |  |
| lnIL-6 | 1.669 | 1.25-2.22 | 0.0004* |
| lnLDH | 3.792 | 1.55-9.28 | 0.0035* |
| lnTnI | 2.02 | 1.58-2.58 | 0.0000* |
| lnFerritin | 2.81 | 1.66-4.76 | 0.0001* |

\*Logistic regression: $p < 0.01$ shows statistical significance.

**Table S4.** Diagnoses in a sub-group of 28 patients presenting with more than one neurological disease concomitant with COVID-19.

|  | Demen<br>tia | Epilep<br>sy | Parkins<br>on<br>Disease | Multiple<br>Sclerosis | Polyneurop<br>a<br>thy | Myasthe<br>nia | SA<br>H | Migrain<br>es | CNS<br>Tum<br>or | Oth<br>er |
| --- | --- | --- | --- | --- | --- | --- | --- | --- | --- | --- |
| Dementia | x | 1 | 4 | x | 1 | 0 | 0 | x | x | 1 |
| Epilepsy | 0 | x | 0 | 1 | 0 | 0 | 0 | 1 | 4 | 1 |
| Parkinson<br>Dis. | 0 | 0 | x | 0 | 1 | 0 | 0 | x | 0 | 1 |
| M. Sclerosis | 0 | 0 | 0 | x | 0 | 0 | 0 | 0 | 0 | 0 |
| Polyneurop<br>a<br>thy | 0 | 0 | 0 | 0 | x | 0 | 0 | 0 | 0 | 0 |
| Myasthenia | 0 | 0 | 0 | 0 | 0 | X | 0 | 0 | 0 | 0 |
| SAH | 0 | 0 | 0 | 0 | 0 | 0 | x | 0 | 0 | 0 |
| Migraines | 0 | 0 | 0 | 0 | 0 | 0 | 0 | x | 0 | 0 |
| CNS Tumor | 0 | 0 | 0 | 0 | 0 | 0 | 0 | 0 | x | 0 |
| Ischemic<br>Stroke | 5 | 5 | 3 | 1 | 0 | 0 | 0 | 0 | 0 | 0 |

